# Epidemiology of Reopening in the COVID-19 Pandemic in the United States, Europe and Asia

**DOI:** 10.1101/2020.08.05.20168757

**Authors:** Weiqi Zhang, Alina Oltean, Scott Nichols, Fuad Odeh, Fei Zhong

**Affiliations:** Merck Holding (China), Shanghai, China, a business of Merck KGaA, Darmstadt, Germany; MilliporeSigma, St. Louis, Missouri, a business of Merck KGaA, Darmstadt, Germany

**Keywords:** COVID-19, Reopening, Age, non-pharmaceutical interventions, hospitalization, United States, Europe, Asia, interrupted time series analysis

## Abstract

Since the discovery of the novel coronavirus (SARS-CoV-2), COVID-19 has become a global healthcare and economic crisis. The United States (US) and Europe exhibited wide impacts from the virus with more than six million cases by the time of our analysis. To inhibit spread, stay-at-home orders and other non-pharmaceutical interventions (NPIs) were instituted. Beginning late April 2020, some US states, European, and Asian countries lifted restrictions and started the reopening phases. In this study, the changes of confirmed cases, hospitalizations, and deaths were analyzed after reopening for 11 countries and 40 US states using an interrupted time series analysis. Additionally, the distribution of these categories was further analyzed by age due to the known increased risk in elderly patients. Reopening had varied effects on COVID-19 cases depending on the region. Recent increases in cases did not fully translate into increased deaths. Eight countries had increased cases after reopening while only two countries showed the same trend in deaths. In the US, 30 states had observed increases in cases while only seven observed increased deaths. In addition, we found that states with later reopening dates were more likely to have significant decreases in cases, hospitalizations, and deaths. Furthermore, age distributions through time were analyzed in relation to COVID-19 in the US. Younger age groups typically had an increased share of cases after reopening.

## Introduction

The novel coronavirus was first discovered in Hubei, China and rapidly spread across the country. On March 11^th^, 2020, the World Health Organization (WHO) declared a global pandemic for the new coronavirus disease (COVID-19)^1^. As of July 20^th^, 2020, there have been >14 million confirmed cases and >597 thousand deaths globally reported by WHO. Among all countries, the coronavirus spread rapidly within the US since its first 100^th^ death was reported on March 16, 2020, and it has reached >3.5 million cases and >137 thousand deaths^2^.

In response to COVID-19, different non-pharmaceutical interventions (NPIs) were implemented across countries worldwide. On Jan. 23^rd^, China put Wuhan, the center of the outbreak, on strict lockdown with significant economic repercussions. Residents were required to stay at home and all public transportation was suspended. Soon after, some Asian countries (e.g., South Korea and Singapore) executed travel restrictions and/or quarantine measures^3^. Beginning mid-March, European countries started to implement country-wide orders to close schools and non-essential businesses, restrict gatherings, limit international travel, and issue stay-at-home orders^4^. On the other hand, the US had a heterogeneous response to COVID-19 that was managed at local municipality and state levels. Most stay-at-home orders became effective from March 21^5^. In the early months of the COVID-19 pandemic, the mobility of retail, recreation, grocery, pharmacy, park, public transit station, and workplace locations were noticeably decreased^6^. The effectiveness of the NPIs in absence of a vaccine for COVID-19 was studied in multiple countries, and researchers found that combinations of NPIs may have great impact on reducing viral transmission^7^. It should be also noted that intense social distancing control such as lockdown was particularly effective^8,9^.

However, nationwide lockdown is unsustainable in the long-term due to negative socioeconomic impacts. It was uncertain how and when to reopen safely. Generally, a gradual and staged release strategy in consideration of local situations was recommended rather than a simple “on-off” strategy^10,11^. Optimal results were achieved by targeted NPIs for different age groups^12^, yet researchers had contrary opinions for school reopening. In some cases, children were urged to go back to school^13^ since they seemed unlikely to become infected^14^ or spread the infection^15^, but in other reports, reopening schools was predicted to likely lead to an increase in COVID-19 cases such as in France^16^ suggesting to delay school reopening^17^.

Age and underlying conditions were among the most significant factors affecting the outcome of COVID-19 from an early report in China^18^. About 80% of deaths in the US occurred in patients with age older than 65^19^, even though this age group is only 16.5% of the US population^20^. The risk for hospitalizations and deaths were 6 and 12 times higher, respectively, for COVID-19 patients with underlying diseases^21^. While less likely to be fatal, medical vulnerability for young adults to develop severe illness was 32% according to a recent study^22^.

Since late April 2020, most states in the US eased restrictions and reopened gradually. Dramatic case increases in the US were observed and reported^23^ leading some states to pause or reverse the reopening process.^24^ Beginning in May, some European and Asian countries cancelled stay-at-home orders. Daily new cases in parts of Europe were stable,^25^ while Germany and parts of Eastern Europe saw cases increase.^26^ This raised the question whether increases in cases after reopening had led to severe outcomes and which of the age groups were mostly affected.

Herein, we retrieved time series data for confirmed COVID-19 cases, hospitalizations, and deaths from publicly available data. Interrupted time series analysis was used to evaluate for significant trend changes after reopening in 40 US states and 11 European and Asian countries. To get a deeper insight, detailed time series data by age were also obtained to analyze which age groups had increased share in cases, hospitalizations, deaths after reopening. This analysis was the first attempt to provide a comprehensive understanding of how various countries and states performed following reopening and to introduce insights into effects that age group infection shares affected observed outcomes.

## Methods

### Data collection and selection

All data were obtained as of July 13^th^, 2020. Reopening dates for US states were collected from the New York Times, as well as paused or reversed reopening status^27^. Forty states had state-wide stay-at-home orders and at least 26 days after reopening (14 days + 12 days incubation period, illustrated below) to ensure enough data for time point analysis. The dataset for cumulative case and death time series was collected from the New York Times^28^ and then calculated as daily new cases and deaths. Current hospitalization data for all selected states except Alabama, Florida, and Kansas were obtained from the University of Minnesota COVID-19 hospitalization tracking project^29^. Alabama^30^, Florida^31^, and Kansas^32^ daily new hospitalizations were collected separately. Hospitalization data for four states, including Georgia, Tennessee, New York, Hawaii, were not available and therefore not included in the analysis.

Reopening dates for other countries were obtained from the Oxford coronavirus government response tracker^33^, by extracting the date of stay-at-home requirement reversal (when indicator C6 changed from non-zero to zero). Eleven European and Asian countries with at least 10,000 cumulative cases were selected using the same criteria as US states. Case and death data were obtained for European and Asian countries from John Hopkins University^34^, and then daily new cases and deaths were calculated from the dataset.

The detailed case, hospitalization, and death data by age were collected daily from government dashboards for the past several months and combined to obtain the time series data^35,36,37,38,39,40,41,42,43,44,45,46,47^. Massachusetts data were collected separately from their official public health website^48^.

### Determination of time period

Five days for case and 12 days for hospitalization and death changes were excluded following the suspension of stay-at-home orders to account for any change in NPIs to take effect^49^. The final effective reopening date, defined as the reopening date plus incubation period of either five or 12 days, was used in statistical analysis to analyze each country and state. Time interval before reopening was selected to be the same number of days after reopening.

### Statistical analysis

An interrupted time series analysis was used to test trend changes for the daily new cases, current hospitalizations, and daily new deaths before and after reopening^50^. The analysis detected the observed variable changes over time. Such analysis has been widely used to evaluate the effectiveness of health interventions implemented at a specific time and has also been used in COVID-19 research^51^. The Poisson Regression model was applied without losing generality. Any negative value in daily new cases/deaths, due to inaccuracy in the data source, was replaced by zero. A P value for the slope coefficient (β) indicating the slope change after the intervention was obtained for each country/state. Significant changes were examined at *P*<0.05 threshold. An indication of increase, stable, or decrease in the different categories was defined as a significant slope change or mean difference by two-tailed Student t-test. A positive slope coefficient after reopening indicates increase trend, and a negative slope coefficient indicates inversely. For states with average daily new deaths less than five, the trend was marked as stable since it was not meaningful to assess so few deaths.

For analysis by age, the average share change for each age group was determined as the difference in percentage of daily new cases, hospitalizations, or daily new deaths before and after the effective reopening date.

## Results

Eight out of 11 countries (Austria, Slovenia, Japan, Romania, France, Serbia, Belgium, Spain) had significant increases in daily new cases after reopening whereas the other three countries (Germany, United Arab Emirates, Poland) showed a reduction in daily new cases (Table 1). The daily new cases of Austria, Germany, Slovenia, Japan and Belgium went through the peak and greatly dropped before reopening, although Slovenia and Japan had potential to reverse the trend afterwards (Supplementary Figure 1). The daily new cases of Romania and Serbia increased rapidly and even surpassed levels prior to reopening. Surprisingly, the daily new cases of the United Arab Emirates increased before reopening and then dropped after the reopening. In the daily new death analysis, only two of 11 countries (Romania, Serbia) showed an increase after reopening. Two countries (Slovenia, United Arab Emirates) had no significant change with average daily deaths less than five, and the other seven countries (Austria, Germany, Japan, Poland, France, Belgium, Spain) showed a significant reduction (Table 1 and Supplementary Figure 2).

**Table 1.**
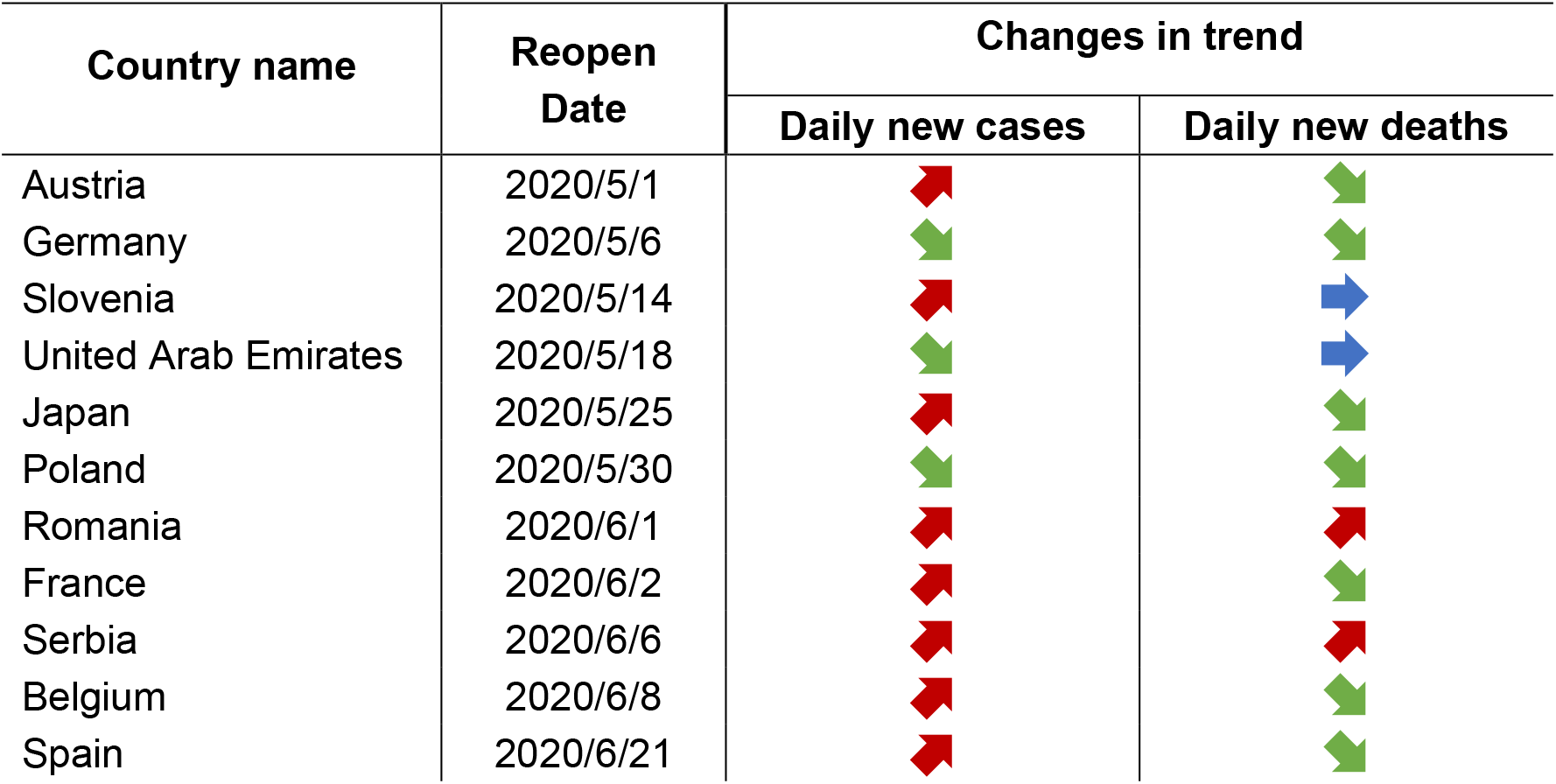
Trends before and after reopening of 11 European and Asian. Changes in trend in “daily new cases” and “daily new deaths” marked with up, flat, or down arrows indicated an increasing, stable, or decreasing trend after reopening in each category, respectively. The increasing and decreasing trends were determined by significant slope change by interrupted time series analysis or significant differences by two-tailed Student’s t-test.

Among 40 states, 30 states had significantly increased daily new cases after reopening (Table 2). Ten states (Colorado, Rhode Island, Maryland, Minnesota, Massachusetts, Connecticut, New York, Maine, New Jersey, New Hampshire) had decreased daily new cases (Supplementary Figure 3). However, only seven states observed significant increases in daily new deaths. Nineteen states showed a significant decrease in deaths, while the remaining 14 states showed non-significant changes in daily death numbers after reopening from COVID-19 (Supplementary Figure 4). Changes in hospitalizations exhibited mixed results following reopening with 17 states significantly increasing hospitalizations and the other 19 states significantly decreasing hospitalizations (Supplementary Figure 5).

**Table 2.**
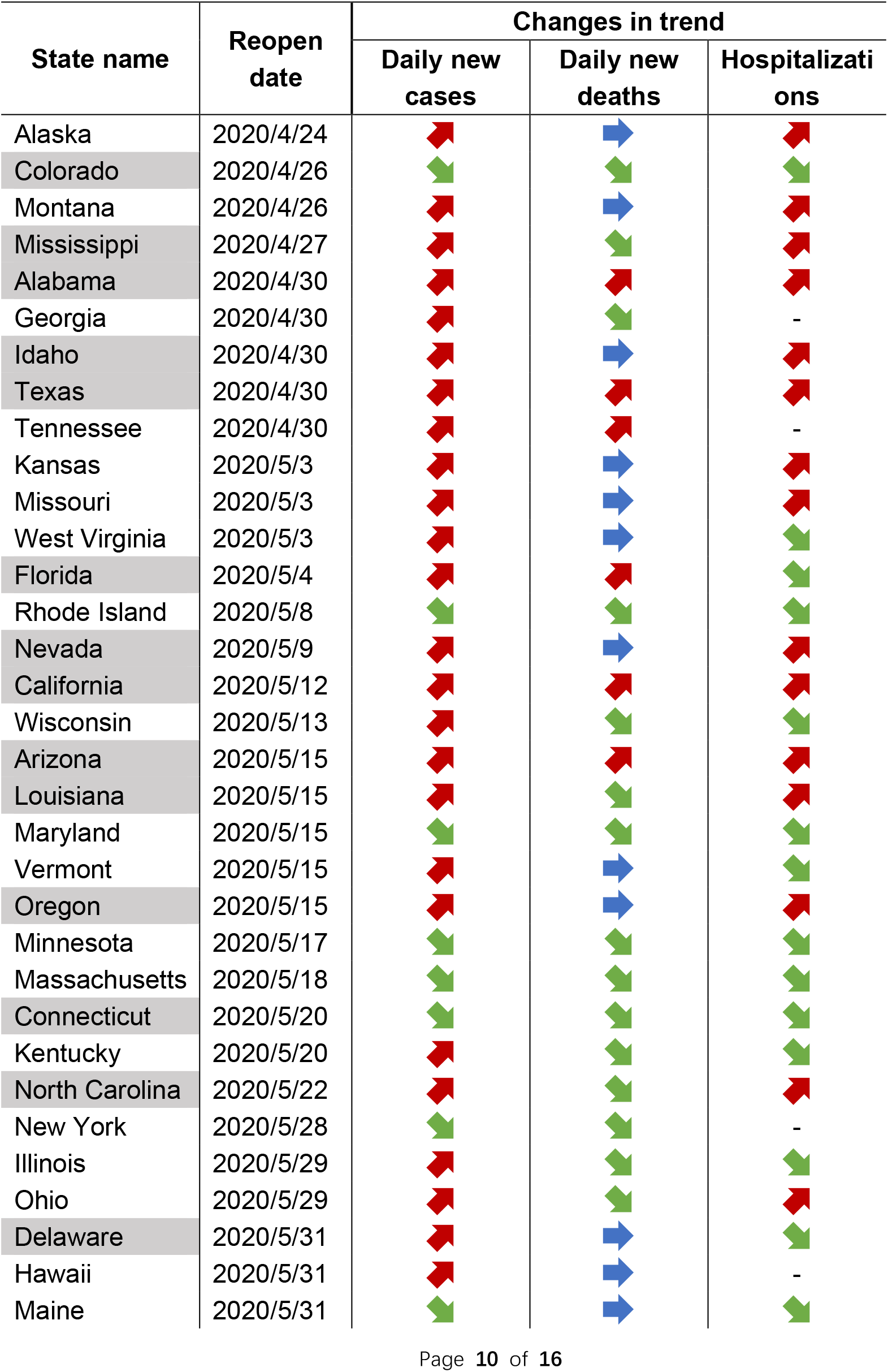
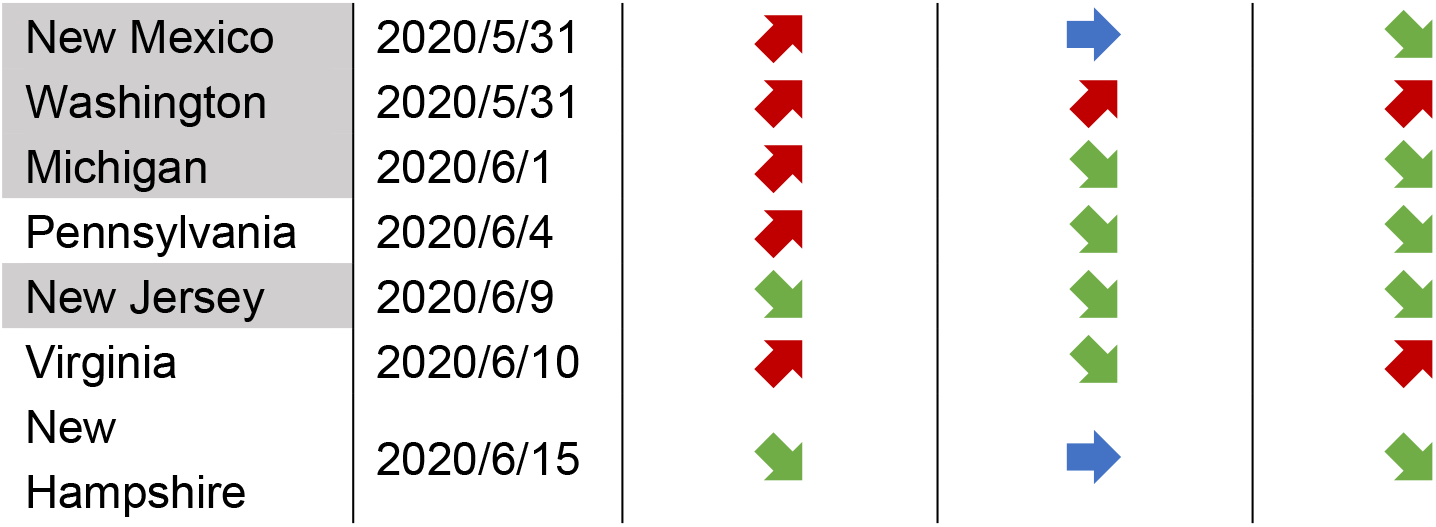
Trends before and after reopening of 40 US states. States with grey background indicated paused or reversed reopening. Changes in trend in “daily new cases”, “daily new deaths” and “hospitalizations” marked with up, flat, or down arrows indicated an increasing, stable, or decreasing trend after reopening in each category, respectively. The increasing and decreasing trends were determined by significant slope change by interrupted time series analysis or significant differences by two-tailed Student’s t-test.

Four countries and 15 US states showed consistent trends in both daily cases and deaths following reopening. While with respect to the combination of cases and hospitalizations, 26 of 36 states with hospitalization data displayed the same trend in both categories (Table 1-2). Before reopening, daily new deaths in most countries were in line with daily new cases; however, only Germany and Romania had a consistent decreasing trend for cases and deaths after reopening (Supplementary Figure 1-2). Eight states (Colorado, Rhode Island, Maryland, Minnesota, Massachusetts, Connecticut, New York, New Jersey) had decreasing trend in both daily new cases and deaths (Supplementary Figure 3-4). When focusing on states sorted by reopening date, it appeared that a delay in reopening date resulted in a higher likelihood of positive outcome (i.e., decreasing daily new cases) (Table 3). The percentage of states with significantly reduced infection rates after reopening was increased from 11.7% with an early reopening date (before May 15^th^) to 34.8% with a late reopening date (after May 15^th^). The results also showed that 60.9% of later-reopening states (14 out of 23 states) have seen a significant decrease in their average daily new deaths in comparison to 29.4% of early-reopening states (5 out of 17 states). The same trend was also observed with hospitalizations.

**Table 3.** Summary of percentage of states in the two reopening periods with increasing or decreasing trends in each category. Sum of decrease and increase may be less than 100% due to stable trends following reopening.

| Reopen date range | Change in new daily cases |  | Change in new daily deaths |  | Change in current hospitalizations |  |
| --- | --- | --- | --- | --- | --- | --- |
|  | Decrease | Increase | Decrease | Increase | Decrease | Increase |
| 4/24-5/14 | 11.7% | 88.2% | 29.4% | 29.4% | 33.3% | 66.7% |
| 5/15-6/15 | 34.8% | 65.2% | 60.9% | 8.7% | 66.7% | 33.3% |

To get deeper insight, we also analyzed whether the age distributions in COVID-19 data had changed after reopening. Age group definitions varied between states, but general trends were still observable. The case share distributed relatively even across age groups, while middle-aged people around 40 to 65 were the largest contributor to hospitalizations, and the elderly aged 60 and over made up most of the COVID-19 deaths (Supplementary Figure 6-8). All 13 states with data showed increased share in younger age groups less than 40 in daily new cases after reopening (Figure 1). The share of hospitalizations in younger age groups for five available states was also increased slightly. However, we did not observe the same trend for daily new deaths.

**Figure 1.**
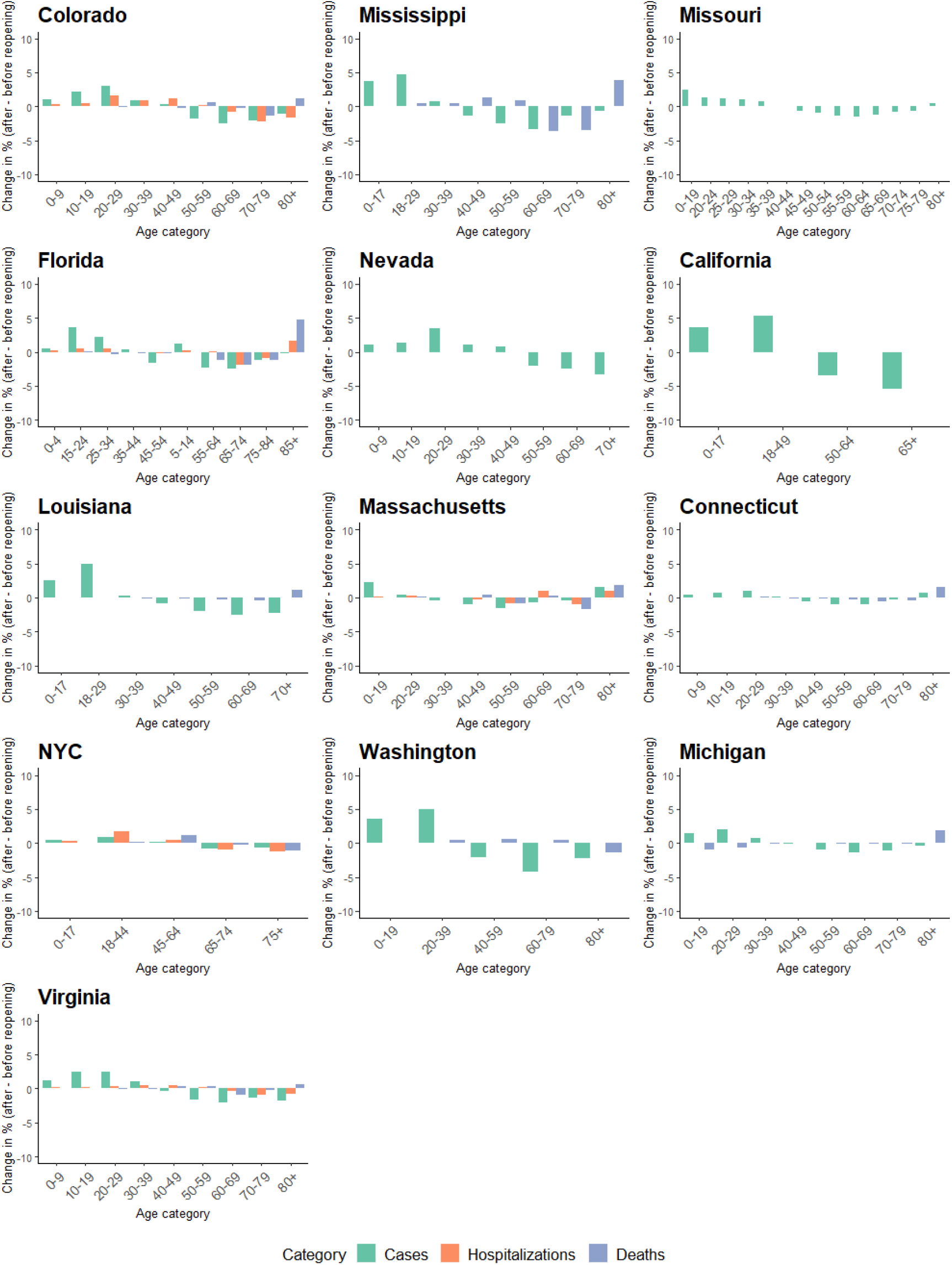
Average share change of age groups on cases, hospitalizations, and deaths before and after reopening in 13 states or regions. States are ordered by reopening date. Death data were available for 10 states and hospitalization data were available for five states.

## Discussion

In this study, interrupted time series analysis was performed to detect significant trend changes in daily new cases, current hospitalizations, and daily new deaths after reopening. The data was collected from publicly available resources with resultant data quality depending on local or regional reporting of the information on COVID-19. Overall in the US, changes in daily new cases and hospitalizations were largely consistent (26 of 36 states). Better outcomes were observed in states with later reopening dates. On the contrary, changes in daily new cases did not necessarily lead to the same changes in daily new deaths. It is not clear why the increase in cases and hospitalizations would not also result in an increase in deaths; however, more time may be needed to observe an increase in deaths due to a lag between case detection and death certificates, especially for those states opened in June^52^. We have not seen any states with increased daily new deaths from our last analysis in June; while in July, deaths in seven states began to increase. Increased testing availability, resulting in more detection of milder cases, could also be a reason for the inconsistent pattern between cases and deaths^53^. The observed increased share of daily new cases in younger age groups also suggests that the average age of COVID-19 is getting younger; these younger people are less likely to suffer severe outcomes from the disease. Alternatively, hospitals and health care providers may now be able to provide better treatment to COVID-19 patients, leading to improved patient survival. The results in European and Asian countries share some similarities with the trends in the US. Some countries had increased daily new cases, but this was not always reflected in daily new deaths. However, European and Asian countries had reopened for a relatively shorter time (8 of 11 reopened after May 15^th^). Other changes may have not manifested yet, and complications, both geographically and culturally, could impact potential differences.

When comparing COVID-19 data between age categories, the case share increases in younger age groups is of particular interest. Less guarded social contacts after reopening may be the most relevant factor, and many news reports are attributing increased case load to young people participating in social activities^54,55^. While younger people have a lower risk of death, they can still transmit the virus to others and potentially impose a great risk to the elderly.

In order to reopen states according to the White House, three conditions should be observed as follows: (1) downward influenza-like illness symptoms, (2) downward COVID-19 cases, and (3) enough hospital resources^56^. State leaders need to illustrate that COVID-19 is no longer spreading uncontrolled. In addition, sufficient testing and hospitalization capacities should be ensured to track and isolate infected people. Unfortunately, many states reopened without meeting the criteria and showed worse trendlines^57^. As of July 20^th^, only four states (Arizona, Delaware, Maine, New Jersey) were trending better in the last 14 days, and the national average of estimated hospital bed availability is less than 40%^58,59^. Concerns have been expressed whether there would be new coronavirus outbreaks following potentially premature reopening of regions. There is no formal definition of a second wave, yet it is possible, especially in US, as several states have reached new highs for daily new cases. Many states such as California and Florida reversed the decision of reopening and chose to close the state again in response to the dramatic increase in cases.

China observed a potential second wave after 56 days without a domestic case in Beijing on June 11^th 60^. In seven days, 158 new cases were confirmed since the first detected case in Xinfadi market and spread to other cities^61^. Closure of related markets, mass testing, tracing and tracking of the cases at community level, centralized hospital resources, and tightened travel control were all performed in one week^62^. Due to a fast response, the small outbreak managed to be sustained within Beijing. The changes on human life from COVID-19 could be long-term^63^. Fast and prepared regional response, including sufficient testing, contact tracing and tracking, might be the best chance to control the potential spread. Spain approved a “new normality” decree to include safety measures to avoid a second wave of infections. Usage of face masks, personal protection measures, and a safe social distance of 1.5 meters were recommended at any public place^64^. Therefore, these NPI measures could be essential to public health before an effective vaccine becomes available in the market.

This study analyzed the impact of reopening in several regions using real-world retrospective COVID-19 related data. Our results found an inconsistency between the trends in cases and deaths. We also concluded that a delay in reopening and postponing the lifting of NPIs resulted in a greater likelihood of decreasing cases, hospitalizations, and deaths. Furthermore, younger people aged less than 40 years old contributed to an increased share of cases among all age groups after several states reopened.

## Data Availability

The data that support the findings of this study are available on request

